# Transmission of SARS-CoV-2 by inhalation of respiratory aerosol in the Skagit Valley Chorale superspreading event

**DOI:** 10.1101/2020.06.15.20132027

**Authors:** Shelly L. Miller, William W Nazaroff, Jose L. Jimenez, Atze Boerstra, Giorgio Buonanno, Stephanie J. Dancer, Jarek Kurnitski, Linsey C. Marr, Lidia Morawska, Catherine Noakes

## Abstract

During the 2020 COVID-19 pandemic, an outbreak occurred following attendance of a symptomatic index case at a regular weekly rehearsal on 10 March of the Skagit Valley Chorale (SVC). After that rehearsal, 53 members of the SVC among 61 in attendance were confirmed or strongly suspected to have contracted COVID-19 and two died. Transmission by the airborne route is likely. It is vital to identify features of cases such as this so as to better understand the factors that promote superspreading events. Based on a conditional assumption that transmission during this outbreak was by inhalation of respiratory aerosol, we use the available evidence to infer the emission rate of airborne infectious quanta from the primary source. We also explore how the risk of infection would vary with several influential factors: the rates of removal of respiratory aerosol by ventilation; deposition onto surfaces; and viral decay. The results indicate an emission rate of the order of a thousand quanta per hour (mean [interquartile range] for this event = 970 [680-1190] quanta per hour) and demonstrate that the risk of infection is modulated by ventilation conditions, occupant density, and duration of shared presence with an infectious individual.

**Practical Implications:**

- During respiratory disease pandemics, group singing indoors should be discouraged or at a minimum carefully managed as singing can generate large amounts of airborne virus (quanta) if any of the singers is infected.
- Ventilation requirements for spaces that are used for singing (e.g., buildings for religious services and rehearsal/performance) should be reconsidered in light of the potential for airborne transmission of infectious diseases.
- Meetings of choirs and other kinds of singing groups during pandemics should only be in spaces that are equipped with a warning system of insufficient ventilation which may be detected with CO_2_ “traffic light” monitors.
- Systems that combine the functions heating and ventilation (or cooling and ventilation) should be provided with a disclaimer saying “do not shut this system off when people are using the room; turning off the system will also shut down fresh air supply, which can lead to the spread of airborne infections.”

## Introduction

SARS-CoV-2 was first reported in China at the end of 2019 and rapidly spread to the rest of the world over the subsequent months. Evidence from laboratory studies has shown that the SARS-CoV-2 virus can remain infectious while airborne for extended periods.^1,2^ The virus has been detected by PCR in the air in several healthcare environments.^3-9^ Researchers have reported values for the SARS-CoV-2 viral load in the mouth that span an extraordinarily broad range: 10^2^ and 10^11^ copies per mL of respiratory fluid.^10-12^ Viral loads vary over the course of the disease, tending to peak at the onset of symptoms.

Airborne transmission is now strongly suspected to play a significant role in superspreading events (SSEs) under certain conditions.^13^ SSEs occur when a large number of secondary transmissions are produced early in an outbreak and transmission is sustained in later stages.^14^ Some people release respiratory aerosol at an order of magnitude greater rate than their peers and might contribute to superspreading events.^15^ The very broad range of viral loads in respiratory fluids may also be an important factor influencing SSE. An infectious respiratory aerosol is a collection of pathogen-laden particles in air emitted during respiring activities of an infected individual. ^16^

In this paper, we first estimate the infectious quanta emission rate during a choir rehearsal that has been identified as a superspreading event. Quanta are used to represent infectious respiratory aerosol when the actual viral dose in the aerosol and the human dose-response required to cause infection are unknown.^17,18^ We then explore the sensitivity of the secondary attack rate of infection to the loss rate of airborne virus, whether by ventilation, deposition onto surface, or biological decay.

## Case Study

An SSE occurred in Skagit Valley, Washington, USA.^19^ When the Skagit Valley Chorale (SVC) met on the evening of March 10, 2020, one person attending the rehearsal had cold-like symptoms that had developed three days earlier; that individual subsequently tested positive for COVID-19. This person is considered the “index case.” At the time of the rehearsal the Skagit County Health Department was not recommending widespread closure of public venues or public events. They were recommending that those 60 y of age and older should avoid large public gatherings. Choral members were told by the director in an email to not attend on March 10 if they were sick with *any kind of symptoms* or if they had concerns.

At the time of the rehearsal, there were no known COVID-19 cases in Skagit Valley County, nor were any closures in effect. The day after the rehearsal on March 11, the governor of Washington recommended physical distancing and no large group meetings in three other nearby counties. Before detecting the cluster on March 17, Skagit county had developed seven COVID-19 cases.

The SVC has 122 members, but only 61 attended rehearsal on March 10, amid concerns about COVID-19 transmission. Precautions were taken during rehearsal, including the use of hand sanitizer, no hugging or handshakes.^20^

Some members began experiencing illness from March 11 to March 15. The timing of these potential secondary infections is consistent with what is known about the temporal dynamics of virus shedding and the serial interval for COVID-19.^21^ Among the 61 attendees at the rehearsal, 53 cases in total were subsequently identified including the index case, with 33 confirmed through positive COVID-19 tests and 20 unconfirmed but probable secondary cases based on symptoms and timing. Accounting for the one presumed index case, the secondary infection attack rate is thus in the range 32/60 to 52/60, or 53-87%.

The chorale met in the Fellowship Hall of a church in Mount Vernon, Skagit County. A seating chart obtained through personal communication showed the layout of participants among 120 chairs plus the position of the choir director and piano accompanist. Although the chart cannot be reproduced because of privacy concerns,^19^ a centrally important point for interpreting the cause of transmission is that the cases were broadly distributed throughout the room with no clear spatial pattern.

The rehearsal started at 6:30 pm. The SVC rehearsed in a single group in the Fellowship Hall for 45 minutes, then split into two approximately equal-sized groups for 45 minutes. One group, mostly male singers, went to practice around a piano in a different room of the church, while a second group stayed in the Fellowship Hall. After practicing separately, and following a 10-minute break, the members reconvened in the Fellowship Hall for another 50 minutes, until 9 pm. During the split session, those who remained in the Fellowship Hall occupied about half of the space.

Limited information is available about the heating and ventilating system; what was learned from personal communications is summarized here. The Fellowship Hall is heated by a relatively new commercial forced-air furnace. Three supply air registers are situated 2.4 m above the floor on one wall with a single return on an adjacent wall, just above the floor (∼0.15 m). Someone in the front office reportedly turned on the heating system prior to the rehearsal to warm the space, and the thermostat was set to 20 °C (68 °F). It was about 7 °C (45 °F) outside, so the heating was on at the start of the rehearsal, but with so many people in the room, it did not need to stay on to maintain a comfortable temperature. During the entire rehearsal no exterior doors were open. It is not known whether the forced-air furnace fan operated (only) under thermostatic control or whether it ran continuously. The furnace is installed with both outside make-up and combustion air, but it is not known how much outside make-up air was supplied that evening. The furnace is also outfitted with a MERV 11 filter, which has a single-pass efficiency of ≥ 30-65% for airborne particles of diameter 1 µm or larger.^22,23^

## Modeling Airborne Infection Risk

Inhalation of respiratory aerosol most likely dominated infection transmission during this event, as other modes of transmission are unlikely to account for the high secondary attack rate. For example, it seems infeasible that all attendees touched the same surface(s) as the index case. Furthermore, rehearsal attendees expressed that they had taken great care to minimize contact transmission (personal communication). There is no evidence to suggest that more than one person was infected at the time of the rehearsal. The index case would have spent extended time within a few meters of only a small proportion of the rehearsal attendees. Other close contact events that extend to a high proportion of the attendees would have been brief and incidental. Consequently, we believe it likely that shared air in the Fellowship Hall, combined with high emissions of respiratory aerosol from singing, were important contributing factors.

On the basis of the available information about this event, a modeling effort was undertaken with two goals. The first goal was to estimate an average quanta emission rate that is consistent with the evidence. The calculation proceeds in two steps: determining the average airborne quanta concentration from the reported secondary infection attack rate, and then evaluating the emission rate that would have produced the inferred average concentration. The second modeling goal was to explore how a change in the loss rates, for example owing to improved ventilation and filtration, would have altered the infection risk. In pursuing both goals, the modeling effort uses an idealization of the more complex real situation, in part because some key data are lacking. A similar approach has been used in other studies to explore airborne infection risk in indoor environments.^17,24^

The model of infection risk due to airborne transmission is based on the Wells-Riley formulation,^25,26^ as amended by Gammaitoni and Nucci.^27^ In applying this approach, these assumptions are made: i) there is one infectious individual who emits SARS-CoV-2 quanta at a constant rate throughout the event, ii) there is no prior source of quanta in the space, iii) the latent period of the disease is longer than the time scale of the model, iv) the infectious respiratory aerosol quickly becomes evenly distributed throughout the room air, and v) infectious quanta are removed by first-order processes reflecting the sum of ventilation, filtration, deposition, and inactivation. The assumption that the indoor environment can be modeled as well-mixed is substantiated in this case by the broad spatial distribution of secondary infections among the rehearsal participants. In epidemic modelling, where the aim is to assess the spread of the disease in the community, it is impossible to specify geometries, ventilation efficiency, and the locations of the infectious sources in each microenvironment. Therefore, adopting the well-mixed assumption is generally more reasonable than hypothesizing about specific patterns of emissions, airflow and removal processes.^28^ This distinctive superspreading event, occurring in an enclosed community facility, with indoor space shared for a specified period of time, offers a unique opportunity to examine a range of physical parameters that influence the eventual outcome.

The modeled probability of infection (*p*) is related to the number of quanta inhaled (*n*) according to equation (1):^26^

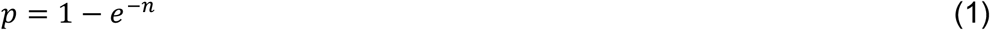

Equation (1) is used to estimate the average quanta concentration during the practice. The airborne quanta concentration increases with time from an initial value of zero following a “one minus exponential” form, which is the standard dynamic response of a well-mixed indoor volume to a constant input source. The time-average quanta concentration (*C*_avg_, q m^-3^) is the quanta inhaled divided by the volume of air breathed. The volume of air breathed (m^3^) is equal to the duration of the event (*D*, h) multiplied by the volumetric breathing rate of rehearsal participants (*Q*_b_, m^3^ h^-1^).

A well-mixed material balance model for the room (equation (2)) is applied next to relate the quanta concentration, *C* (quanta per m^3^), to the emission rate, *E* (quanta per h):

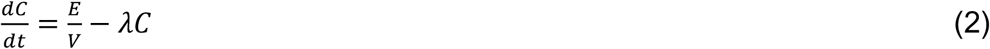

Here *V* = volume of the rehearsal hall (m^3^) and *λ* = first-order loss rate coefficient for quanta (h^-1^) due to the summed effects of ventilation (*λ*_*v*_), deposition onto surfaces (*λ*_*dep*_), and virus decay (*k*).^29^ Assuming the quanta concentration is 0 at the beginning of the rehearsal, equation (2) is solved and the average concentration determined as follows:

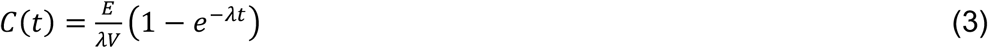

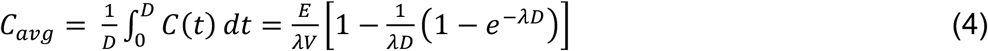

Here, *t* = time (h). Equation (4) is rearranged to solve for the emission rate, *E*:

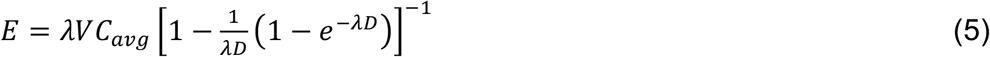

A Monte Carlo simulation was run (*N* = 1000) to estimate *E* for the superspreading event given a range of input values. The unknown parameters (*p, Q*_b_, *λ*_*v*_, *λ*_*dep*_, *k*) were specified as probabilistic using uniform distributions bounded by specified upper and lower limits. These parameters were assumed to be uncorrelated.

The ranges of the uncertain model parameter values explored in the Monte Carlo simulation are summarized in Table 1. Constant values were used for the volume of the Fellowship Hall and the rehearsal duration.

**Table 1.**
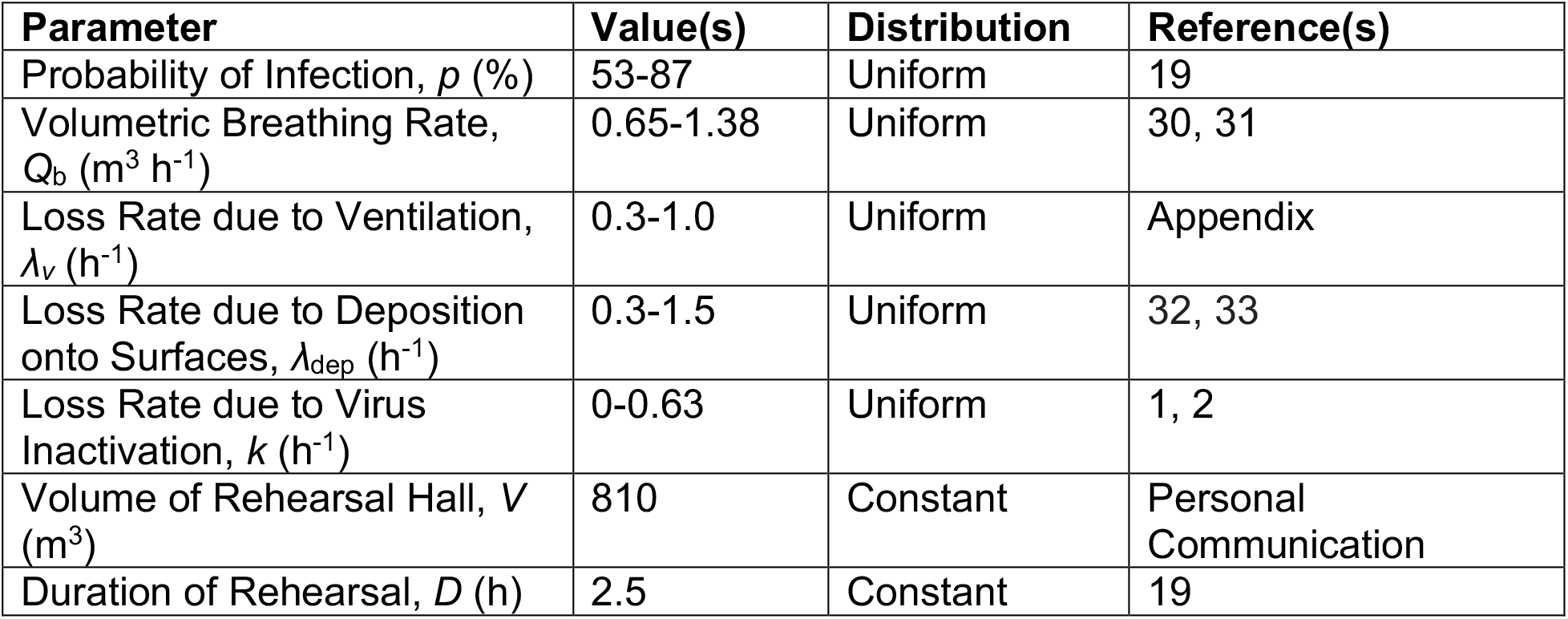
Parametric Values used in the Monte Carlo Simulation for Estimating *E*

Volumetric inhalation rates of singers have been reported by Binazzi et al.^31^ to be in the range 0.22-1.0 m^3^ h^-1^ and by Adams et al.^30^ to be 1.38 m^3^ h^-1^. SARS-CoV-2 was found in air samples in two size ranges: 0.5-1 µm and > 2.5 µm.^7^ The surface deposition loss rate range was based on data from Thatcher et al.^33^ and Diapouli et al.^32^ The range of values for virus decay is based on two sources: Fears et al.^1^ showed no decay in virus-containing aerosol for 16 hours at 53% RH, whereas van Doremalen et al.^2^ estimated the half-life of airborne SARS-CoV-2 is 1.1 h, which equates to a decay rate of 0.63 h^-1^. The loss rate due to ventilation is likely to have been in the range from 0.3 to 1 h^-1^ (see Appendix).

## Results

The mean (± standard deviation) inferred emission rate was *E* = 970 (± 390) quanta per h. Additional statistics for the distribution of *E* from the Monte Carlo simulation are as follows: geometric mean = 900 q h^-1^; geometric standard deviation = 1.5; 10^th^, 25^th^, 50^th^, 75^th^ and 90^th^ percentiles: 550, 680, 910, 1180, 1510 q h^-1^.

The emission rate was derived based on an assumption of one index case. It is plausible that more than one person attending the rehearsal was infectious, given that the disease was diagnosed in some of the singers soon after the March 10 rehearsal. If this was the case then our emission rate would be the sum of emission rates from each infectious individual. However, the average incubation time for this case was ∼3-4 days, which is comparable to literature reports, making the presence of additional index cases less likely.

Quanta emission rates for influenza have been reported to be in the range 15-128 quanta h^-1^;^17,34^ for measles: 5,580 q h^-1^;^35^ and for tuberculosis: 1.25 to 30,840 q h^-1^ (the high value attributed to intubation).^36^ The quanta for SARS transmission in a hospital and in an elementary school was estimated to be 28 q h^-1^.^37^ A forward model was used to estimate a large range of estimated quanta emission rates for SARS-COV-2, depending on activity level and respiratory activity: 10.5-1030 quanta h^-1^.^24^

To explore the influence of changing the loss rate on the probability of infection, we performed sensitivity simulations in which we varied the loss rate. In these simulations, we used the mean emission rate of *E* = 970 q h^-1^ and a constant volumetric breathing rate of *Q*_b_ = 1.0 m^3^ h^-1^. If *λ* is systematically increased by some combination of increased ventilation, deposition, filtration, and inactivation loss rates, how would the probability of infection decrease? We also explored what would happen if the emission rate was set at the 10^th^ and 90^th^ percentile values from the Monte Carlo simulation.

Using the model equations above with *λ* ranging from 0.6 to 12 h^-1^, the percentage of the rehearsal participants infected is determined. The results are plotted in Figure 1.

**Figure 1.**
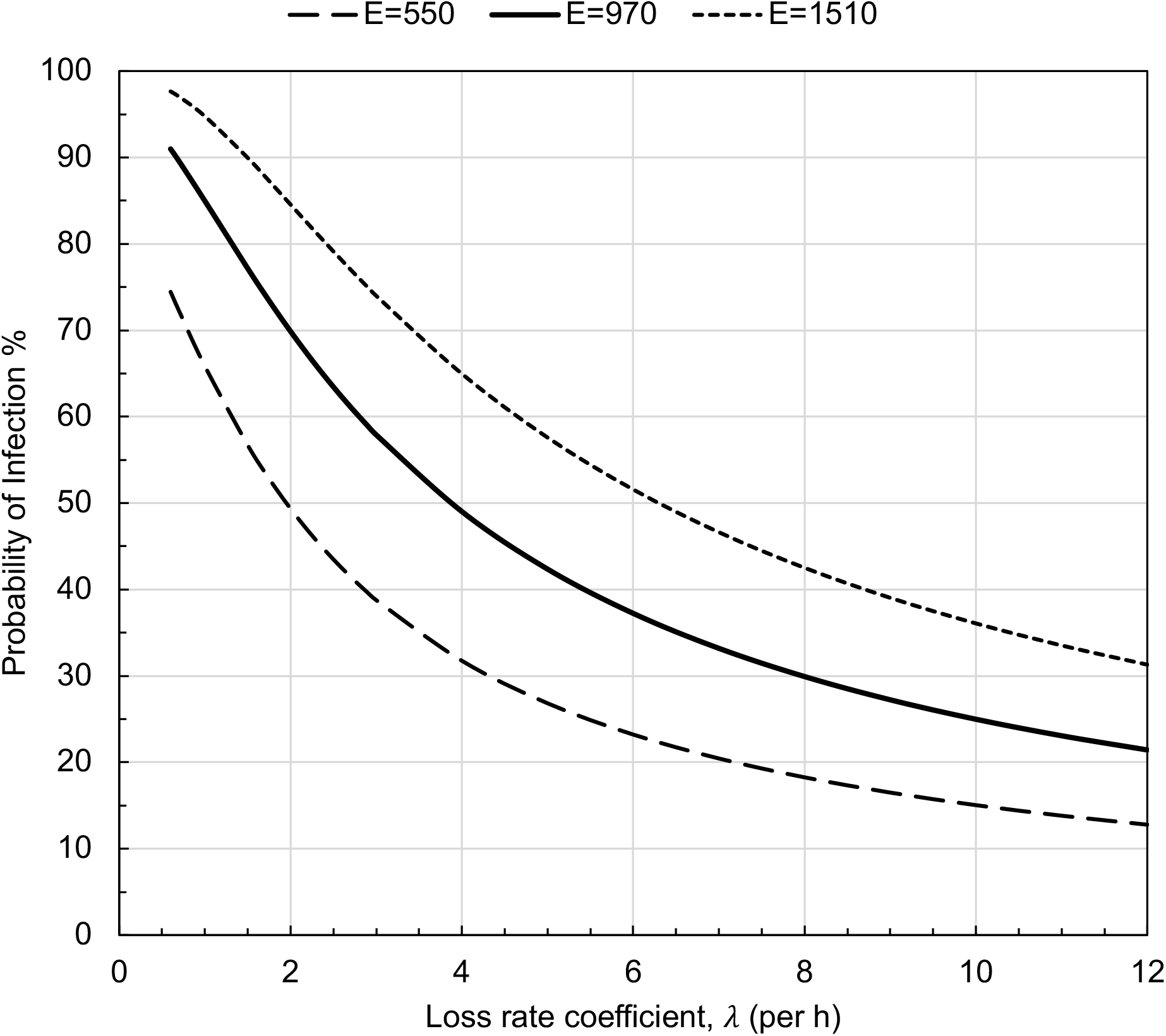
Probability of infection for each rehearsal participant as a function of loss rates for varying airborne quanta emission rates (E, q h^-1^). Infection probability is plotted for the predicted mean emission rate (970 q h^-1^) and the 10^th^and 90^th^ percentile emission rates (550 and 1510 q h^-1^, respectively.) A rehearsal duration of 2.5 hours, an indoor volume of 810 m^3^ and a volumetric breathing rate of 1.0 m^3^ h^-1^ were assumed.

A key point displayed in Figure 1 is that, for the mean value *E* = 970 q h^-1^, increasing the loss rate coefficient from a nominal baseline value of 0.6 h^-1^ to 5 h^-1^ would reduce the probability of infection by a factor greater than two, from 91% to 42%. For the full range of loss rates plotted in Figure 1, the infection risks spans a factor of four: from 91% to 21%.

We also explored how changing the duration of the event would impact the probability of infection as a function of loss rate. Again, we use the mean emission rate of 970 q h^-1^ and a volumetric breathing rate of 1.0 m^3^ h^-1^. For durations ranging from 0.5 to 2.5 hours, and λ ranging from 0.6 to 12 h^-1^, the predicted percentage infected ranged broadly, from 4% to 91%. The results are plotted in Figure 2.

**Figure 2.**
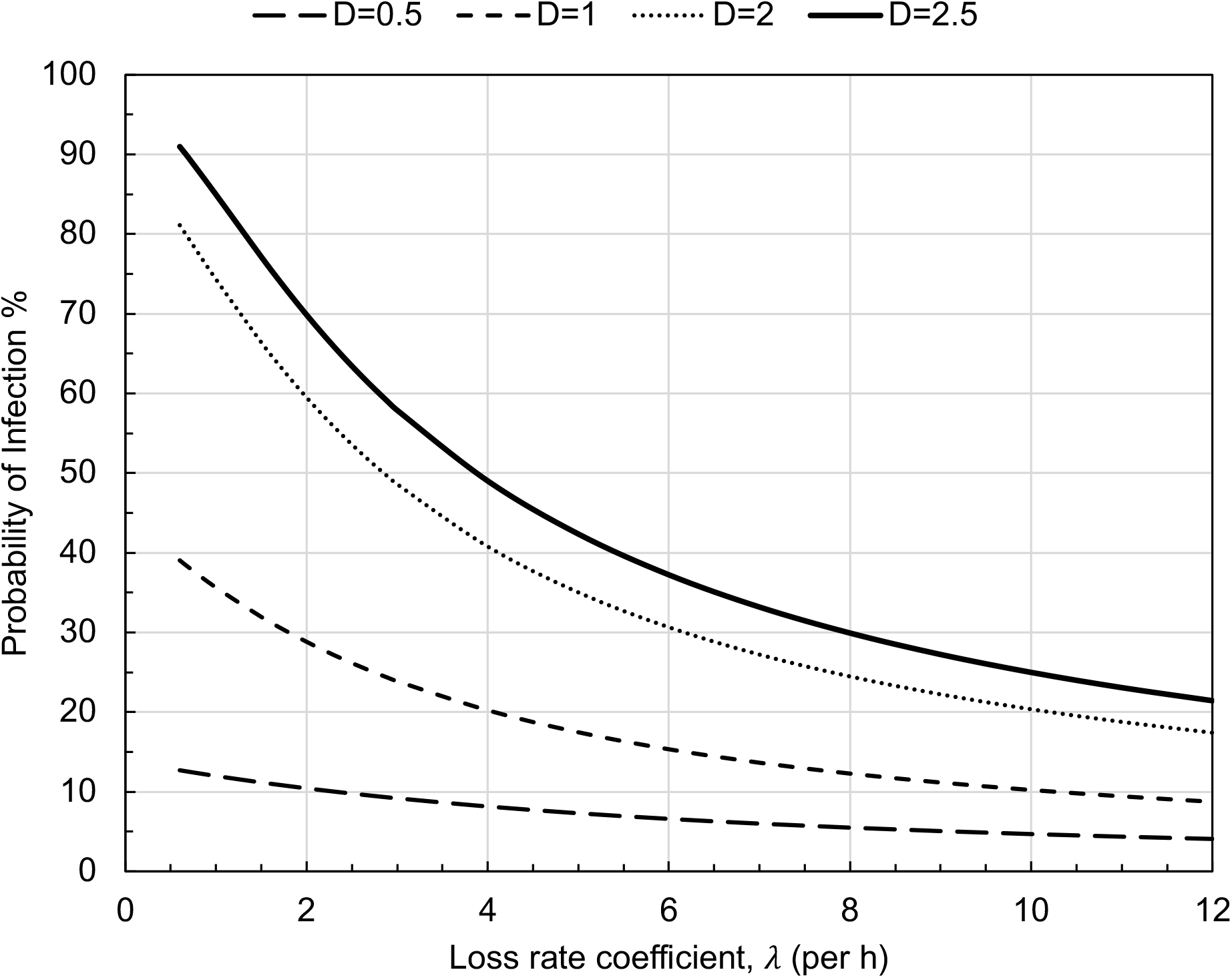
Probability of infection as a function of loss rates for varying event duration (D, h). A mean emission rate (970 q h^-1^) and constant volumetric breathing rates of 1.0 m^3^ h^-1^ were assumed.

## Discussion

Growing evidence supports a view that inhaling respiratory aerosol is an important route for transmission of SARS-CoV-2 under certain conditions. At the time of the chorale rehearsal on 10 March 2020, because of emerging concern about SARS-CoV-2, person-to-person contact and touching of surfaces was consciously limited. The risk of widespread transmission owing to close contact would seem to be low in this event, considering that there is believed to have been only one index case who would have been seated in proximity to only a small proportion of the other chorale members. If transmission by close contact and/or fomites were indeed the dominant modes of transmission, then the secondary attack rate should have been much smaller than the observed range of 53-87%. We would also expect to see the secondary cases predominantly among those in closer proximity to the index case rather than distributed broadly throughout the room. Given the circumstances of the rehearsal, such a high secondary attack rate by the close-contact route would have necessitated effective transmission based largely on brief proximate encounters. That interpretation of the high attack rate in this event seems much less probable than the alternative explanation, i.e. that inhalation of infectious respiratory aerosol from “shared air” was the leading mode of transmission.

Literature evidence suggests that singing could have been a contributing factor to the high secondary attack rate compared to other common indoor activities. The rate of aerosol emission during vocal activities increases with the loudness of the sound.^15^ A study of respiratory emissions also found higher emission rates of respiratory droplets to be associated with more extensive vocalization.^38^ Outbreaks of tuberculosis, a disease known to be transmitted via inhalation, have been linked to singing.^39-41^ At the time this article is being written, there have been additional media reports of COVID-19 outbreaks associated with choirs. Cases with high secondary attack rates have been reported in the Netherlands, Austria, Canada, Germany, England, South Korea, and Spain.^42,43^

Loudon and Roberts^44^ characterized respiratory aerosol emitted during talking, singing and coughing. They reported that “fewer droplets were expelled during singing than during talking, but a higher proportion of them were in the smaller size range. The percentage of droplets still airborne as droplet nuclei after a 30-minute settling period were 35.7, 6.4, and 48.9 for singing, talking, and coughing, respectively.”

The inferred emission rate of 970 quanta h^-1^ is plausible given observations of airborne concentrations of SARS-CoV-2 in hospitals. The highest concentrations reported averaged 3000 ± 2700 viral RNA copies m^-3^ across 18 measurements in Nebraska^9^ and 2600 ± 1000 viral RNA copies m^-3^ and across two measurements in Singapore.^3^ In the Singapore study, the highest values were measured on day 5 of illness where a symptomatic patient had a high viral load in their nasopharyngeal swab (Ct value 18.45). If the dominant removal mechanism is ventilation at an average rate of 13 h^-1^ in Nebraska and 12 h^-1^ in Singapore, then these concentrations correspond to emission rates of the order of 10^6^ viral RNA copies h^-1^ from a patient. Typically, only 0.1-1% of viral RNA copies represent an infectious virion for influenza,^45^ so if that value is applicable to SARS-CoV-2, the emission rate would correspond to 1000-10,000 infectious virions emitted per hour; viral load emitted also varied between coughing and breathing/speaking.^45^ Lindsley et al. have shown this effect, too, for infectious influenza virus.^46^

The plausibility of the inferred quanta emission rate can also be demonstrated by combining evidence on respiratory aerosol emissions with viral loads for SARS-CoV-2 in saliva. Concentrations of respiratory aerosol in exhaled breath that are smaller than 10 µm diameter are in the approximate range 1-10 nL m^-3^ for vocalization activities.^38^ For this concentration range, a volumetric breathing rate of 1 m^3^ h^-1^ would produce an emission rate of 1-10 nL h^-1^ of respiratory aerosol. In limited sampling of SARS-CoV-2 in saliva and other respiratory fluids, viral loads as high as 10^11^ viral RNA copies mL^-1^ have been reported.^10,11,12,50^ At 10 nL h^-1^, a viral load in respiratory fluids of 10^11^ RNA copies mL^-1^ (= 10^5^ copies nL^-1^) would lead to the emission of 10^6^ viral RNA copies per h, which would be of order 1000 quanta h^-1^ assuming an equivalence of 1 quanta to 1000 viral RNA copies.

This modeling analysis has explored the very probable situation in which transmission by inhaling respiratory aerosol that were released during singing caused a large COVID-19 outbreak. Accumulating evidence points to these factors being important for increasing the risk of airborne transmission indoors: high occupancy, long duration, loud vocalization, and poor ventilation.

In the domain of indoor environmental quality control, the first and best measure is generally to minimize indoor emissions.^47^ Because we are not yet able to identify individuals who are highly infectious and therefore are potential superspreaders, effective source control can not be so well practiced, short of suspending high-risk indoor events. The simulation results presented here show that the risk of secondary infections can be substantially reduced although not practically eliminated through a combination of increasing removal rates and by limiting the duration of indoor activities. The high ventilation rate in the hospital settings combined with other controls such as use of isolation rooms and effective PPE is likely to mitigate transmission from a high viral shedder in the healthcare environment.^3,9^ In the many community indoor spaces not dedicated to infection control, controlling airborne diseases transmission remains a great challenge during this pandemic. Ventilation rates corresponding to current standards would allow occupancy duration of only about 0.5 h for an infection risk level below 10% for a such high emission activity as investigated here. Indoor environmental quality control measures available to improve conditions include enhanced ventilation, mechanical filtration, and germicidal ultraviolet disinfection.^48,49^ Widespread application of effective indoor environment controls could help limit the extent of superspreading events and therefore contribute to slowing the pandemic spread.

## Data Availability

Data available on request

## Appendix

### Ventilation Rate Estimates

The ventilation rate was estimated assuming that the HVAC fan was not operating during the rehearsal and that the metabolic energy generated by the SVC rehearsal attendees was sufficient to maintain a comfortable temperature without supplemental heating. For conditions of metabolic activity at 1.2 met, clothing insulation of 1.0 clo, at 22 °C (71 °F), the metabolic heat generation per occupant is 78 W. ^51^ Assuming that half of the metabolic energy goes to continuously heat the room air (with the other half lost through the building envelope by conduction and to heat storage), then each occupant would contribute 39 W to the ventilation air. Given the reported difference between indoor and outdoor temperature (23 °F = 13 K) and the heat capacity of air (1 J g^-1^ K^-1^), one can derive the ventilation rate to be 39 W person^-1^ ÷ 13 J g^-1^ = 3 g/s per person. At a density of 1.2 g/L, the resulting ventilation flow rate would be 2.5 L/s per person. For a room volume of 810 m^3^ with 61 occupants, the corresponding air-change rate would be 0.7 per h^-1^. We bracket this estimate by applying an uncertainty of ± 50% so that the modeled range in the Monte Carlo simulation is 0.3-1 per h.

By way of comparison, we have estimated the outdoor air ventilation rate based on the relevant ASHRAE standard combined with information from the mechanical drawings for the rehearsal hall under the assumption that the HVAC fan was on for the event duration. The outdoor make-up air flow specified by ASHRAE Standard 62.1 for places of worship (Table 6.2.2.1) is 2.5 L s^-1^ person^-1^ + 0.3 L s^-1^ per m^2^ of floor area. ^52^ The default occupant density is 120 persons per 100 m^2^ of floor area. The corresponding outdoor air rate per m^2^ of floor area would then be (120/100) × 2.5 + 0.3 = 3.3 L s^-1^ m^-2^. The reported averaging ceiling height for the Fellowship Hall is 4.5 m and the estimated floor area is 180 m^2^. The total ventilation flow rate would therefore be 180 × 3.3 = 594 L s^-1^ = 2100 m^3^ h^-1^, corresponding to an air-change rate of 2.6 h^-1^. Additionally, mechanical drawings of the rehearsal hall show specifications of 3 × 1560 cfm supply registers (indicated to be 8 ft above the floor along one wall). This information indicates that the ventilation system is designed to supply 4700 cfm = 8000 m^3^ h^-1^, which would be a mixture of outdoor air and recirculated indoor air (filtered through a MERV 11 filter). That supply flow rate corresponds to 10 room volumes per hour. Applying the outdoor air flow rate from ASHRAE 62.1, at this overall flow rate, the mix would be about 25% outside air and 75% recirculated air. These 2.5-10 effective air changes per hour are unlikely to have been provided during the rehearsal, based on personal communication received during our investigation.

